# “Populist Attitude and Conspiracist beliefs contribution to the overconfidence about the risk of Covid-19: implications for Preventive Health Behaviors”

**DOI:** 10.1101/2022.01.30.22269992

**Authors:** Giuliani Agnese, Presaghi Fabio

**Author notes:** Correspondence should be addressed to: Agnese Giuliani, Fabio Presaghi.

## Abstract

Populism and Conspiracy beliefs seem to represent the zeitgeist of people depending on fast and simple information retrieved through social media. The Covid-19 emergency has simply catalyzed this process, not without consequences. Supported by literature review, we hypothesize that the higher the populist attitude the higher the tendency in believing in conspiracies, and that both higher populist attitudes and conspiracist beliefs may induce people in underestimating health related risks that may be reflected in a lowered tendency in adopting preventive health behaviors against Covid-19 spread. Data collected during the quarantine (December 2020, March 2021) mainly supported our hypotheses. Results are discussed in accord with the dramatic consequences it may have overconfidence in undermining the adoption of preventive health behaviors.

---

Since the Covid-19 pandemic has begun, we witnessed a reiterated attempt by the main populist leaders all around the world to minimize the severity of the public health crisis. In Italy, the downplaying of the risk of Covid-19 epidemic, carried out by populist opposition parties, such as Lega of Matteo Salvini or Fratelli d’Italia of Giorgia Meloni, was appreciated by a large portion of the Italian population, that is that part of the electorate in search of simplistic solutions to the critical unprecedented situation we’re facing. Furthermore, the search for alternative answers to the “official” ones has led to an increase in the circulation of fake news and conspiracy theories, very often reposted by populist leaders’ official accounts. An emblematic example of this dynamic was the suggestion made by populist leaders, such as Bolsonaro (in Brazil), Trump (in U.S.A.), Salvini and Meloni (in Italy) to use hydroxychloroquine to treat Covid-19 disease, without any scientific evidence of the drug efficacy in this field.

In the present study we hypothesize that populist approach to COVID-19 pandemic along with belief in conspiracy theories may have contributed to undermine the risk of the deceased and may have ultimately interfered with the adoption of safety prescriptions by lay people. Consequently, people may decide to not comply with preventive behaviors, deeming them unnecessary, given their overconfidence about the risk of Covid-19.

## The relation between Populism and Conspiracy Beliefs

Not rarely, the rising of populism rates among the population and the spread of conspiracy theories seem to go hand in hand. From previous definitions it emerges how populists see the élites as “one homogeneous corrupt group that works against the ‘general will’ of the people […] some shadowy forces that continued to hold on to illegitimate powers to undermine the voice of the people” [1]. Interestingly, this narrative is also typical of many conspiracy theories. As Sutton and Douglas [2] argued “[t]o believe in any conspiracy theory is to believe that authorities can be malevolent, that they can conceal their evildoing, and that official explanations for major events may be lies”.

It should come as no surprise that authors who study populism pointed out that politicians of this mold define and portray their opponents as a conspiratorial élite [3, 4, 1] and in cases, openly resort to real conspiracy theories to describe certain events: Albertazzi [5] recalls how far-right parties in Italy often claim that immigrants and refugees are there with the only purpose of creating “unholy coalitions to ruin law-abiding citizens”. Another example is represented by former US President Donald Trump, who was one of the most illustrious exponents of the so-called “birther movement” and encouraging people to believe in the false statement that then candidate Obama to U.S.A. presidency was ineligible as he was not a natural-born citizen of the U.S., as required by Article Two of the American Constitution.

Over time, scholars have tried to identify possible reasons why certain people tend to believe more easily in conspiracy theories: among the personal traits which have been found to play a role in this process, there is also a Manichean vision of reality, which reduces events to a simplistic clash between good and evil [6]. This characteristic is common to both populism and conspiracist mentality [4, 7].

As pointed out by Mark Fenster [8], so-called conspiracy theories proliferate in an environment in which there is “the extreme – indeed, ultimate – skepticism of the political sphere by a sector of the population that feels excluded” [8]. Similarly, believing in conspiracy theories necessitates a belief by citizens that politicians are exclusively engaged in illicit activities aimed at deceiving the population and organizing plans to gain global control [8]. This cynical and disillusioned view of the political sphere and its institutions is also clearly associated with the construct of populism and with the strong preference for parties of this ideological and communicative orientation [9]. Finally, Castanho Silva et al. [10] found a significant and positive correlation between conspiracy beliefs and populism, corroborating the link between the two constructs.

## Conspiracy Beliefs, compliance with preventive behaviors and the overconfidence about the risk of Covid-19

What happens when populist parties endorse conspiracy theories in affirming that preventive health behaviors, like those requested for reducing the risk of Covid-19, are just means to take people under control? Maftei and Holman [11] conducted a study in Romania aimed at exploring the relationship between the tendency to believe in conspiracy theories and three different variables, namely the perceived risk of Covid-19 disease, the perception of adequacy of anti-Covid-19 rules established by the government and the personal disposition to follow those rules. Based on previous studies [12, 13], the researchers expected to find a strong relationship between high levels of the variable called “conspiracist beliefs” and a general skepticism towards preventive norms, such as social distancing and the mandatory use of the mask. Results showed a strong positive correlation between the perceived risk and compliance with preventive behaviors. Furthermore, both perceived risk and the adherence to preventive norms were found to be negatively correlated with conspiracist mentality. These findings are in line with previous research, which had highlighted how the endorsement of conspiracy beliefs represent a concrete obstacle for the compliance with preventive treatments and behaviors, and how these beliefs often negatively affect health-related behaviors [14, 15].

## How overconfidence about the risk of Covid-19 may affect compliance with preventive health behaviors?

As it will be more widely discussed in the next section of this paper, we also hypothesized a relation between the perceived severity of Covid-19 disease and people’s willingness to respect preventive behaviors. This relation was also investigated by Plohl and Musil [16]. In particular, the authors’ expectation was that people who were concerned the most about Covid-19 severity would also be more inclined to comply with preventive behaviors. Results confirmed this hypothesis by highlighting a significant positive relation between perceived severity of Covid-19 disease and citizens’ compliance with preventive behaviors. These results are also in line with those obtained in past studies [17, 18, 19].

## Populism, Conspiracy Beliefs and Preventive Behaviors

We have so far described studies in which the conspiracy mentality was taken into consideration as a possible obstacle to the implementation of preventive behaviors. However, despite the strong relation between populist attitude and the tendency to believe in conspiracy theories, also shown by Castanho Silva et al. [10], there are only few studies that have taken into consideration how populism may affect the perception that specific group of people (like conspiracist groups) might have developed about the Covid-19 pandemic. Moreover, the existence of this relation could have an impact on people’s adherence to preventive behaviors by simply altering (i.e., reducing in this case) their confidence in perceived risk. In a study conducted by Stecula and Pickup [20], both populism and the spread of conspiracy theories was taken into consideration, with the aim of investigating their role in how Covid-19 pandemic was perceived by a specific group of citizens. In particular, the authors’ goal was to describe how populism has been fueling the spread of conspiracy theories during the current pandemic crisis, and the impact that this mechanism seems to be having on individuals’ decision to comply with preventive norms against the spread of SARS-CoV-2. Scholars have hypothesized that populist people are more likely to believe in conspiracy theories and are therefore less motivated to respect preventive rules and experts’ recommendations. The study revealed a significant and positive correlation between populism and conspiracy beliefs, which in turn was negatively correlated with individuals’ adherence to preventive behaviors. Although the authors seem to suggest the existence of a mediation effect of populism on preventive behaviors through the effect of conspiracy beliefs, they did not test a mediation model to investigate this hypothesis. Moreover, as far as we know, there does not seem to be any previous study which has explored a possible direct relation between populism and compliance with preventive rules, just as there do not appear to be studies that have hypothesized a positive effect of populism on the increase of overconfidence about the risk of Covid-19 disease. For these reasons, these two possible relations are advanced and tested in a mediation model in the current study.

## Main Hypotheses

Based on literature previously discussed and on the specific context of Covid-19 epidemic, we hypothesize that Populism may affect Conspiracy Beliefs. We believe that the direction of this relationship better describes what has been vehiculated by national and international media, where populist leaders endorsed conspiracist themes to undermine scientific experts who were trying to stress the real risk about Covid-19. This endorsement may have reinforced populist people’s conspiracy beliefs regarding an unspecified ordeal arranged by elites in the attempt to control people. Furthermore, given that both populism and conspiracy beliefs point to undermine the opinion of scientific experts about the risk of Covid-19, both may significantly increase the overconfidence about the risk of the disease by substantially undermining it. Moreover, the increase in the overconfidence about the risk of Covid-19 may be reflected in a lowered tendency to adopt preventive health behaviors, like those prescribed during the Covid-19 emergency (i.e., the quarantine, distancing from others and mask prescriptions).

## Method

### Participants

Given that the hypothesized relationships basically represent a serial mediation with two mediators (beliefs in conspiracy theories and overconfidence about the risk of Covid-19), we estimated the optimal sample size following the procedure described in Schoemann, Boulton and Short [21]. By setting an expected power of .90, a type 1 error rate of .05, and a conservative standardized regression coefficient of .20 for the three hypothesized relationships we obtained an ideal sample size of about 400 units (power 95% Monte Carlo C.I. with 20000 draws and 5000 replications: .93; .95).

A sample of 477 participants (Females *n* = 313) with an average age of about 38 (*SD* = 14) took part in the online study (after giving a written informed consent) that run from December 2020 to March 2021 (i.e., during the quarantine and at the beginning of the availability of the Covid-19 vaccine in Italy). About 86.8% (*n* = 386) of the sample reported to have not been infected by Covid-19 virus and just 10% (*n* = 47) declared to be already vaccinated against Covid-19, while about 32.5% (*n* =155) declared to have the intention to take the vaccine and about 60% (*n* = 286) of the sample preferred to do not respond. Finally, about 51.8% (*n* = 247) of participants declared to be politically left-center oriented, while about 34.2% (*n* = 163) declared to be politically center oriented and the remaining 14.1% (*n* = 67) was politically right-oriented.

### Data availability statement

All data and scripts for replicating all results discussed in the manuscript are available at the following address: https://osf.io/q8fwt/?view_only=59ae52b6c7b540848c13d25ad8201ee3.

### Analysis Strategy

In order to investigate main hypotheses, we run Confirmatory factor Analysis (CFA) for testing factor structures still to be adapted in Italy measures and Structural Equation Model (SEM) for testing the structural paths among latent factors. We run a preliminary check of multivariate normality of items of each factor structures considered in the SEM (populism, conspiracy believes, perceived risk, prevention health behaviors), and we found that in all cases the assumption was violated to some extent (respectively: Anti-Establishment Attitudes *Henze-Zirkler test* = 4.99, *p* < .001; Demand for Sovereign of the People *Henze-Zirkler test* = 7.78, *p* < .001; Belief in Homogeneity of People *Henze-Zirkler test* = 6.53, *p* < .001; Generic Conspiracist Beliefs *Henze-Zirkler test* = 4.60, *p* < .001; Perceived risk *Henze-Zirkler test* = 14.11, *p* < .001; Preventive Health Behaviors *Henze-Zirkler test* = 104.21, *p* < .001). Given these results, we opted for the Diagonally Weighted Least Square (DWLS, [22]) as method of estimation of SEM parameters as these methods have been shown to give consistent parameter estimates when multivariate normality is violated [23]. In order to assess the goodness of model fit, we used other than the normal theory Chi-squared statistics, also absolute fit indices like Root Mean Squared Error of Approximation (RMSEA) and Standardized Root Mean Squared Residual (SRMR), with relative fit indices like Comparative Fit index (CFI) and Non-Normed Fit Index (NNFI or Tucker-Lewis Index). Values of RMSEA and of SRMR below .08, and values of CFI and NFI above .90 are indicative of model good fit [24].

For those measures for which we cannot retrieve a published validation study from literature (in general or adaptation study in our country), we run a Confirmatory factor analysis in order to inspect psychometric properties of the hypothesized factor structure. In these cases, we also estimated reliability indices: Cronbach alpha) [25], and McDonald’s omega [26] for first and the omega L2 for second order factors [26].

All analyses were run with R software [27] and psych package [28]. For multivariate normality check we used the mvn package [29]. For SEM we used the lavaan package [30]. For reliability of second order factors, we used the semTools package [31]. Indirect effects were estimated with *bootstrap* procedure (with 1000 bootstrap samples) as indicated in McKinnon, Lockwood, and Williams [32].

### Procedure and Measures

All participants completed the following list of questionnaires:

Populist Attitude Scale (PAS) developed by Schulz, Muller, Schemer, Wirz, Wettstein and Wirth [33]. The PAS measures three dimensions of Populism attitude on a 5-steps graded responses scale (1 = *strongly disagree*; 5 = *strongly agree*): the Anti-Elitism Attitude (AEA) with 4 items (e.g. “MPs in Parliament very quickly lose touch with ordinary people”, “People like me have no influence on what the government does”); the Demand for Sovereign of the People (DSP) with 4 items (e.g. “The people should be asked whenever important decisions are taken”, “The politicians in Parliament need to follow the will of the people”); and finally the Belief in Homogeneity of People (BHP) with 4 items (e.g., “Ordinary people all pull together”, “Ordinary people are of good and honest character”). Even if the Italian version of PAS may be found in the literature, we did not find a validation study reporting the psychometric properties of PAS-ita, therefore we proceeded to item translation and back-translation by mother tongue experts. We tested the PAS-ita factor structure (three first order factors with one second-order factor) [33] in a Confirmatory Factor Analysis (CFA). Results showed that the model has good fit indices (Χ^*2*^(51) = 73.41, *p* = .022; *RMSEA* = .030, *RMSEA 90% C*.*I*.: .012; .045; *SRMR* = .047; *CFI* = .992; *NNFI* = .990). Completely standardized factor loadings for the three first order factors were all positive and statistically significant and ranging from .452 to .694 for AEA factor, from .638 to .877 for DSP factor and ranging from .573 to .774 for BHP factor. Finally, all the three factors saturated positively and significantly (with estimates ranging from .619 to 7.58) on the second order factor (Populism Attitude). Regarding reliability, the AEA reported lowest values (*Cronbach alpha* = .63; *McDonald’s omega* = .64), while DSP (*Cronbach alpha* = .87; *McDonald’s omega* = .88) and BHP (*Cronbach alpha* = .81; *McDonald’s omega* = .81) reported satisfactory values. Finally, the Populism Attitude second-order factor reported a discrete reliability (*McDonald’s omega L2* = .72).

Generic Conspiracist Beliefs scale (GCB) is a 15 items scale developed by Brotherton, French and Pickering [34]. The GCB assesses 5 highly correlated dimensions (or facets as Authors call them) of conspiracist beliefs: Government Malfeasance (GM, 3 items, e.g. “The government uses people as patsies to hide its involvement in criminal activity”), Extraterrestrial Cover-up (ET, 3 items, e.g. “Evidence of alien contact is being concealed from the public’’), Malevolent global conspiracies (MG, 3 items, e.g. “Certain significant events have been the result of the activity of a small group who secretly manipulate world events’’), Personal Well Being (PW, 3 items, e.g. “Technology with mind-control capacities is used on people without their knowledge”), Control of Information (CI, 3 items, e.g. “Groups of scientists manipulate, fabricate, or suppress evidence in order to deceive the public’’). However, the use of these 5 dimensions is unclear as in Brotherton et al [34] authors develop and refine the 5 factors structure in the first 2 studies, but then in study 3 and 4 (concerning respectively criterion-related and discriminant validity aspects of GCB) the same authors use the GCB as uni-dimensional, leaving the reader a little disappointed on how to use the scale. In any case, we did not find a validation study providing psychometric properties of the GCB in Italy, so we preferred to investigate the factor structure of the GCB after translation and back-translation procedure. In CFA we compared fit indices of two competing factor structures: the uni-dimensional GCB and the five first-order correlated factors. Contrary to our expectation, the uni-dimensional factor structure performed better (Χ^*2*^(90) = 108.43, *p* = .090; *RMSEA* = .021, *RMSEA 90% C*.*I*.: .000; .034; *SRMR* = .065; *CFI* = .998; *NNFI* = .998) than the five factors structure (Χ^*2*^(81) = 1059.35, *p* < .001; *RMSEA* = .159, *RMSEA 90% C*.*I*.: .151; .168; *SRMR* = .163; *CFI* = .990; *NNFI* = .871; the five correlated factors model did not converge. In order to obtain it, we had to fix to zero the latent correlation between PW and CI). The reliability indices of GCB factor were excellent (*Cronbach alpha* = .94; *McDonald’s omega* = .94). The Generic Conspiracist Beliefs score was computed as the sum of the 15 items. The higher the score the higher the tendency in believing in conspiracy.

The Overconfidence about the Risk of Covid-19 (ORC) was assessed with four items (rated on a 5-point rating scale ranging from 1 = *strongly disagree* to 5 = *strongly agree*) appropriately developed for this study. The four items (two positively and two negatively phrased) were as in the following: “Being infected by Covid-19 o some variants won’t have severe consequences for me”, “It can get hard to treat people positive for Covid-19 or its variants [reversed]”, “Being infected by Covid-19 or by its variants can be really risky [reversed]”, “Covid-19 or its variants are not worse than a bad flu”. Cronbach alpha was slightly below the acceptability (*Chronbach alpha*= .65), while McDonald’s omega was satisfactory (*McDonald’s omega* =.74). So as an indicator of the Overconfidence about the Risk of Covid-19 we computed the average score of the four items. Higher scores indicate a higher overconfidence.

Preventive Health behaviors were assessed with the following four items (rated on a 5-point rating scale ranging from 1 = *strongly disagree* to 5 = *strongly agree*) appropriately developed for this study: “I always take the distance from other people in crowded spaces”, “I have reduced social interactions”, “I follow and I have followed all Government prescriptions, including those concerning movements across regions”, “I always wear adequately the mask”. The analysis of reliability resulted in adequate estimates (*Cronbach alpha* = .86; *McDonald’s omega* = .88). As an indicator of the Preventive Health Behaviors, we computed the average score of the four items. So, the higher the score the higher the tendency to endorse preventive health behaviors.

Finally, all participants completed a socio-demographic sheet where they were asked about: gender identification, age, occupation, level of education, political endorsed party (“If we went to elections tomorrow, which party would you vote for, most likely?”), if they had been infected by Covid-19 (“to your knowledge, have you already contracted Covid-19?”), if they got vaccinated or intended to get the vaccine in future (“Have you already received the Covid-19 vaccine?”, “If not yet, will you get vaccinated against Covid-19 when it’s going to be your turn?”) and if they considered themselves as at-risk subjects (“Do you think you have at least one of the diseases that constitute a risk factor for Covid-19? e.g. immunosuppression, ischemic heart disease, stroke, arterial hypertension, diabetes mellitus etc.”).

All procedures and measures included in the study were in accordance with the ethical standards approved by the Department of “Psychology of Development and Socialization Processes” Ethics Committee, which gave approval to our study, and with 1964 Helsinki declaration on ethical standards.

## Results

Table 1 shows descriptive statistics for items and for latent factors considered in the present study.

**Table 1.**
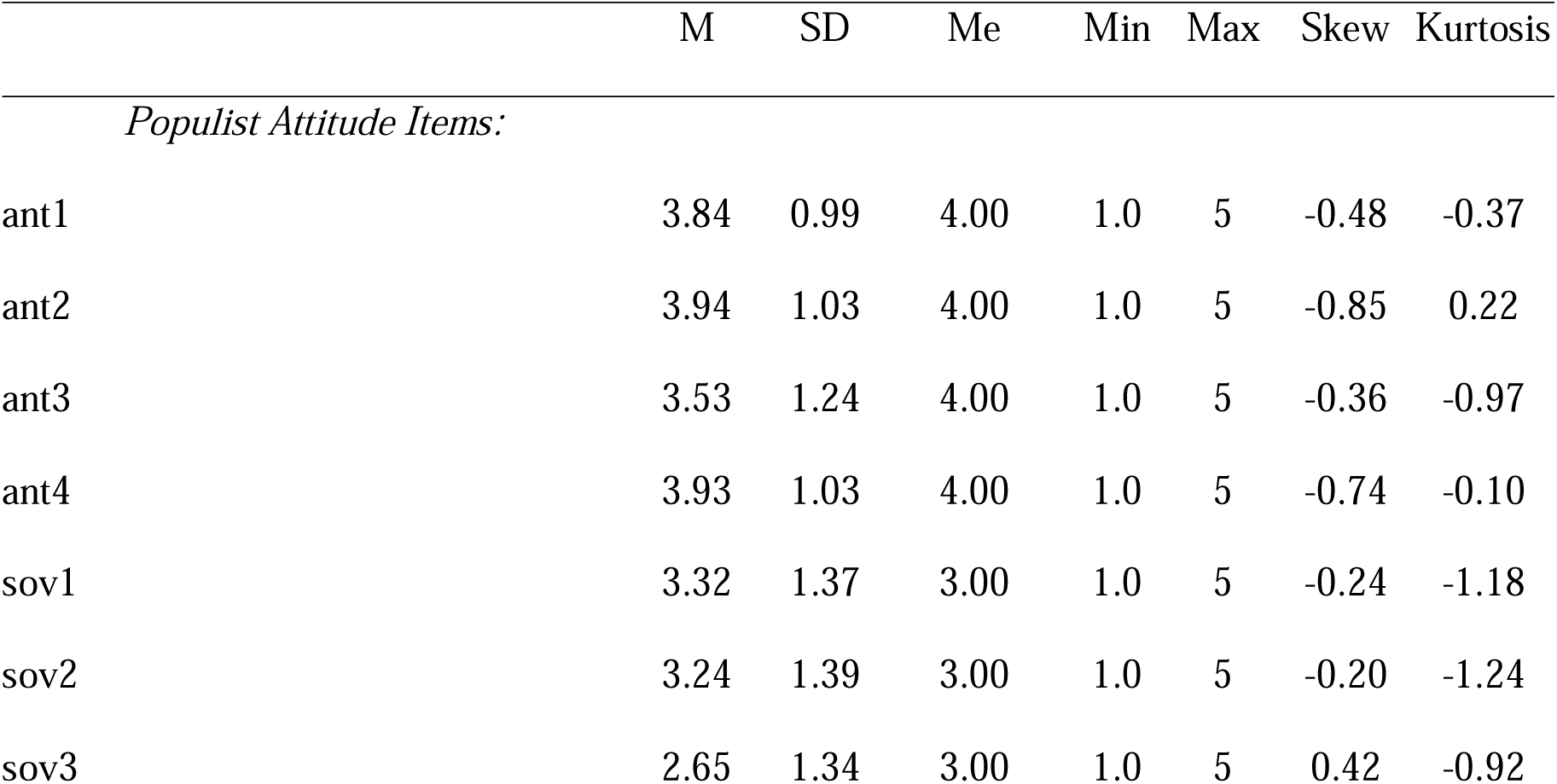
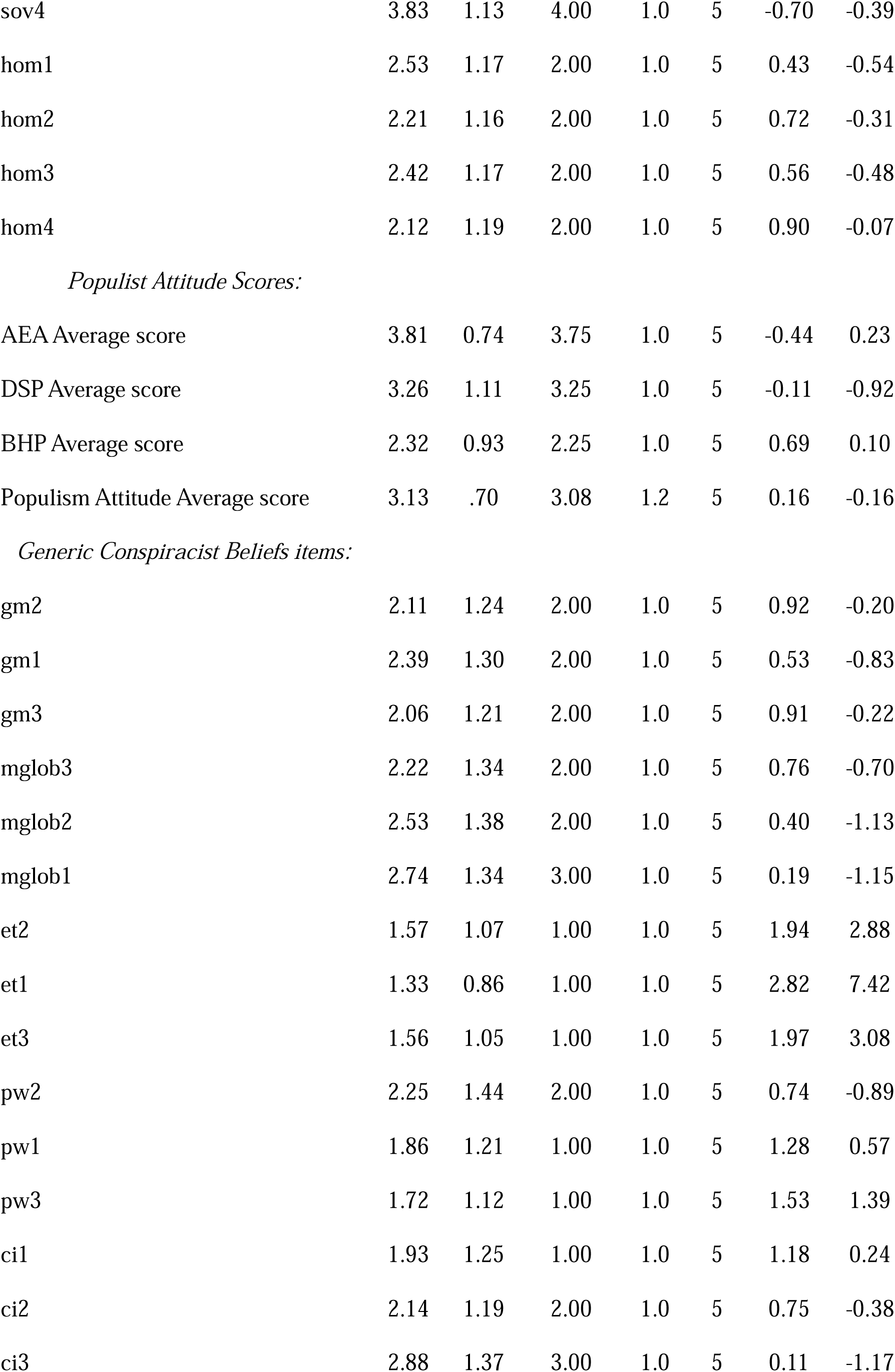
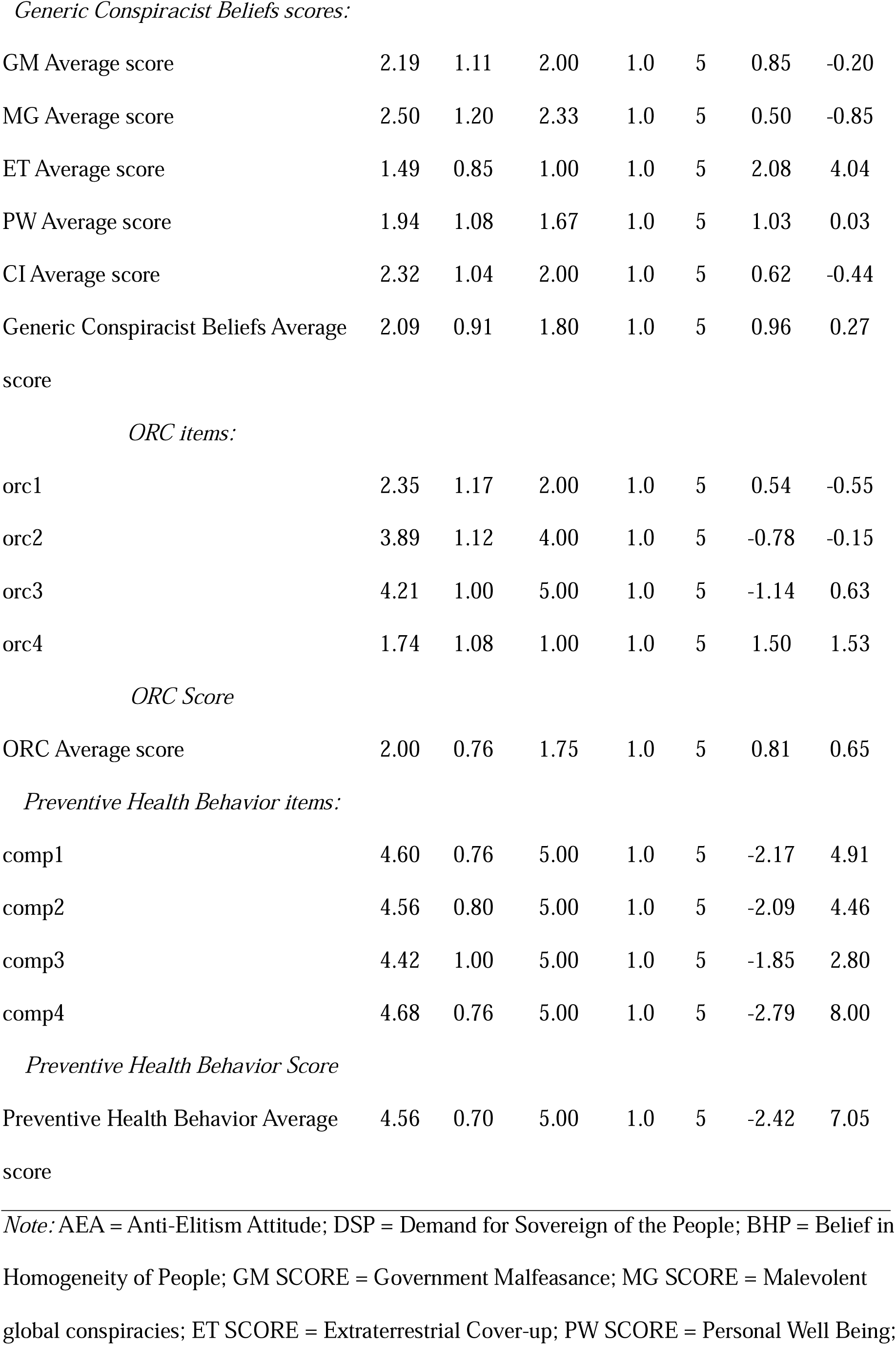
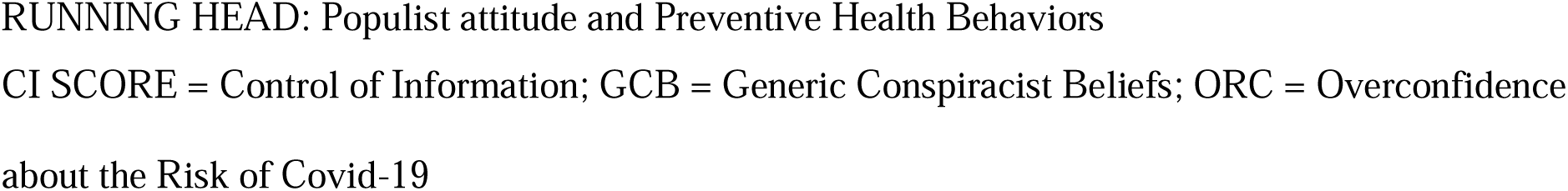
Descriptive statistics (M, SD, Me, Min-Max, Skewness and Kurtosis) for observed and latent factors indicators

*Note:* AEA = Anti-Elitism Attitude; DSP = Demand for Sovereign of the People; BHP = Belief in Homogeneity of People; GM SCORE = Government Malfeasance; MG SCORE = Malevolent global conspiracies; ET SCORE = Extraterrestrial Cover-up; PW SCORE = Personal Well Being; CI SCORE = Control of Information; GCB = Generic Conspiracist Beliefs; ORC = Overconfidence about the Risk of Covid-19

### Structural Equation Model

Results of the Structural Equation analysis showed the hypothesized model has really satisfactory fit indices (Χ^*2*^(81) = 584.37, *p* = .118; *RMSEA* = .012, *RMSEA 90% C*.*I*.: .000; .020; *SRMR* = .054; *CFI* = .998; *NNFI* = .998). Considering the structural paths (table 2, figure 1) we can see that all the three populist attitudes (AEA, DSP and BHP) have significant and positive effects on GCB (respectively: AEA *b* = .246, *se* = .193, *bootstrap 95% C*.*I*.: .274; 1.039; DSP *b* = .237, *se* = .059, *bootstrap 95% C*.*I*.: .094; 0.463; BHP *b* = .256, *se* = .078, *bootstrap 95% C*.*I*.: .156; 0.463). Turning to the prediction of the ORC, we see that none of the populist attitudes have a significant impact (respectively: AEA *b* = -.114, *se* = .099, *bootstrap 95% C*.*I*.: -.329; .067; DSP *b* = .127, *se* = .029, *bootstrap 95% C*.*I*.: -.003; 0.112; BHP *b* = .096, *se* = .044, *bootstrap 95% C*.*I*.: -.033; 0.148), while GCB has a positive and significant effect on Perceived Risk (*b* = .321, *se* = .047, *bootstrap 95% C*.*I*.: .061; .242). Finally, the three populist attitudes were not significant also when considering the prediction of Preventive Health Behaviors (respectively: AEA *b* = .015, *se* = .106, *bootstrap 95% C*.*I*.: -.211; .216; DSP *b* = -.013, *se* = .029, *bootstrap 95% C*.*I*.: -.064; .055; BHP *b* = .091, *se* = .047, *bootstrap 95% C*.*I*.: -.032; .164). However, both GCB (*b* = -.169, *se* = .039, *bootstrap 95% C*.*I*.: -.167; -.012) and ORC (*b* = .537, *se* = .230, *bootstrap 95% C*.*I*.: -1.057; -.414) reported a significant and negative effect. So, to sum up, we found that three populist attitudes are positively associated with GCB, that the GCB is positively associated with the Overconfidence about the Risk of Covid-19 (ORC), and that both GCB and the Overconfidence about the Risk of Covid-19 are negatively associated with Preventive Health Behaviors.

**Table 2.**
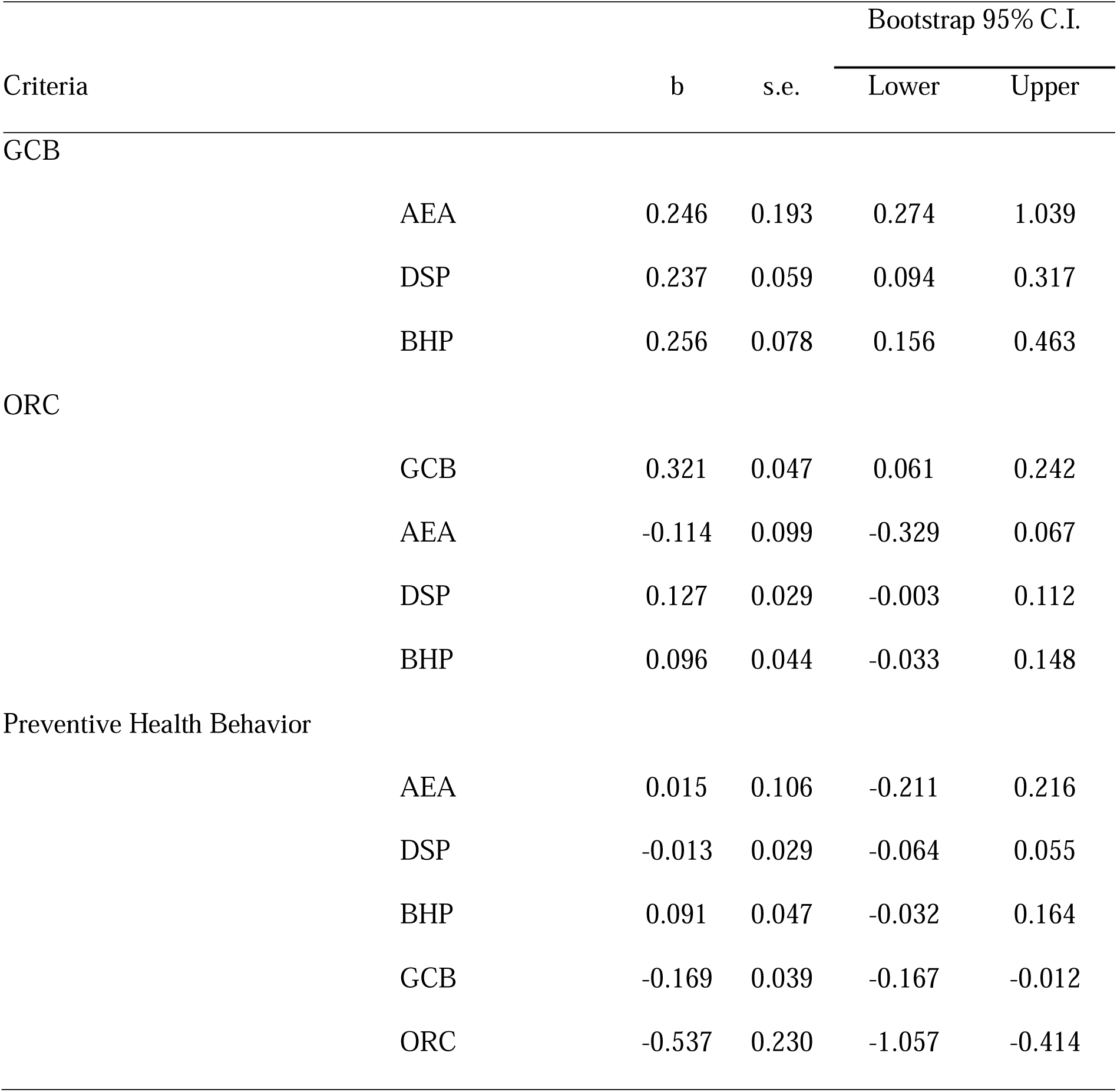
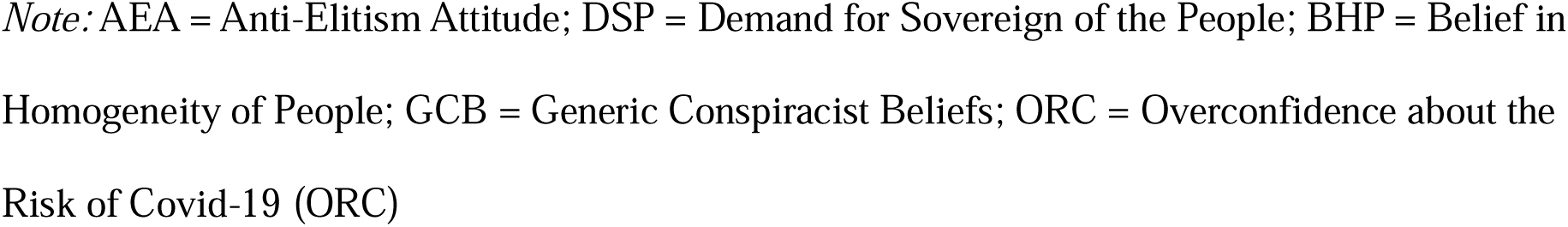
Completely standardized structural coefficients, s.e., and bootstrap 95% C.I. (for non-standardized coefficients)

|  |  |  |  | Bootstrap 95% C.I. |  |
| --- | --- | --- | --- | --- | --- |
| Criteria |  | b | s.e. | Lower | Upper |
| GCB |  |  |  |  |  |
|  | AEA | 0.246 | 0.193 | 0.274 | 1.039 |
|  | DSP | 0.237 | 0.059 | 0.094 | 0.317 |
|  | BHP | 0.256 | 0.078 | 0.156 | 0.463 |
| ORC |  |  |  |  |  |
|  | GCB | 0.321 | 0.047 | 0.061 | 0.242 |
|  | AEA | -0.114 | 0.099 | -0.329 | 0.067 |
|  | DSP | 0.127 | 0.029 | -0.003 | 0.112 |
|  | BHP | 0.096 | 0.044 | -0.033 | 0.148 |
| Preventive Health Behavior |  |  |  |  |  |
|  | AEA | 0.015 | 0.106 | -0.211 | 0.216 |
|  | DSP | -0.013 | 0.029 | -0.064 | 0.055 |
|  | BHP | 0.091 | 0.047 | -0.032 | 0.164 |
|  | GCB | -0.169 | 0.039 | -0.167 | -0.012 |
|  | ORC | -0.537 | 0.230 | -1.057 | -0.414 |

*Note:* AEA = Anti-Elitism Attitude; DSP = Demand for Sovereign of the People; BHP = Belief in Homogeneity of People; GCB = Generic Conspiracist Beliefs; ORC = Overconfidence about the Risk of Covid-19 (ORC)

**Figure 1.**
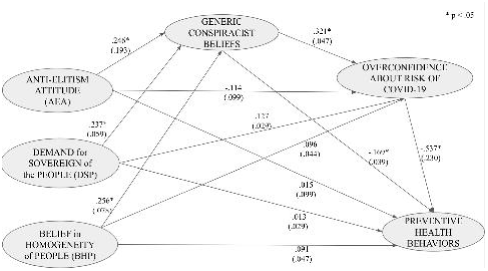
Structural Relationships among Populist attitude, Generic Conspiracist Beliefs, Overconfidence about the Risk of Covid-19 and Preventive Health Behaviors

### Indirect Effects

As can be seen in Table 3, the three populist attitude does not have a significant indirect effect on Preventive Health behaviors through the ORC (respectively: AEA→ORC→Preventive Health Behavior b = .061, se = .062, bootstrap 95% C.I. -.045; .202; DSP→ORC→Preventive Health Behavior b = -.068, se = .020, bootstrap 95% C.I. -.075; .002; BHP→ORC→Preventive Health Behavior b = -.052, se = .028, bootstrap 95% C.I. -.092; .020). However we found a significant and negative indirect effect of populist attitudes on Preventive Health Behaviors through GCB and through ORC (respectively: AEA→GCB→ORC→Preventive Health Behavior b = -.042, se = .024, *bootstrap* 95% C.I. -.114; -.019; DSP→GCB→ORC→Preventive Health Behavior b = -.041, se = .008, *bootstrap* 95% C.I. -.038; -.006; BHP→GCB→ORC→Preventive Health Behavior b = -.044, se = .012, *bootstrap* 95% C.I. -.054; -.010). Therefore, an increase in populist attitude may determine a lowered tendency in adopting Preventive health related behaviors through the increase in conspiracist beliefs, which in turn increase the overconfidence about health-Covid-19 related risk. Pairwise comparisons were considered for contrasting the longest indirect effects. Results (table 3) showed that no significant differences emerged between any couple of indirect effects. Hence all the three longest indirect effects have equal impact on Preventive Health Behaviors.

**Table 3.**
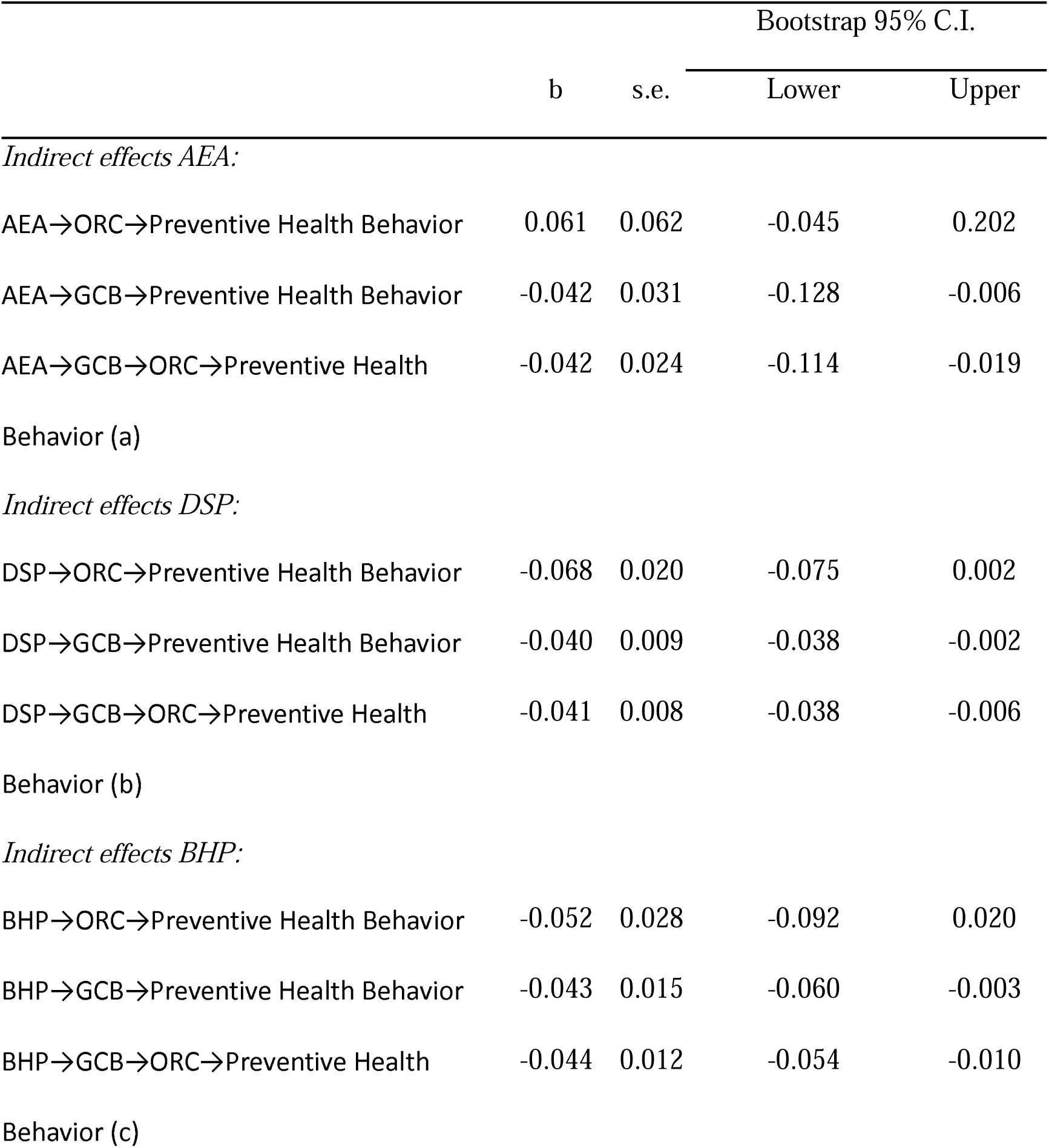
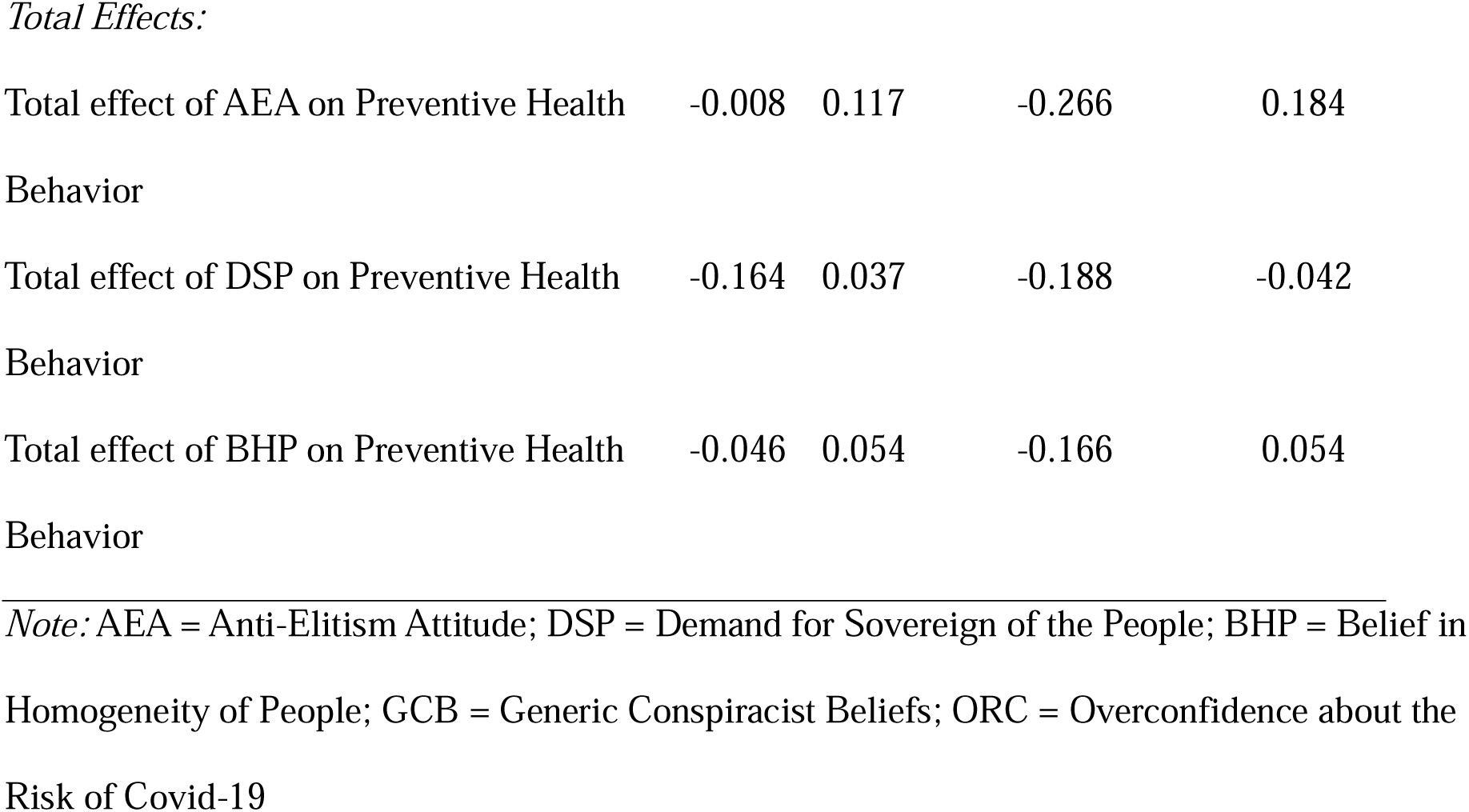
Indirect and Total effects: completely standardized structural coefficients, s.e., and bootstrap 95% C.I.

|  | b | s.e. | Bootstrap 95% C.I. |  |
| --- | --- | --- | --- | --- |
|  |  |  | Lower | Upper |
| *Indirect effects AEA:* |  |  |  |  |
| AEA→ORC→Preventive Health Behavior | 0.061 | 0.062 | -0.045 | 0.202 |
| AEA→GCB→Preventive Health Behavior | -0.042 | 0.031 | -0.128 | -0.006 |
| AEA→GCB→ORC→Preventive Health Behavior (a) | -0.042 | 0.024 | -0.114 | -0.019 |
| *Indirect effects DSP:* |  |  |  |  |
| DSP→ORC→Preventive Health Behavior | -0.068 | 0.020 | -0.075 | 0.002 |
| DSP→GCB→Preventive Health Behavior | -0.040 | 0.009 | -0.038 | -0.002 |
| DSP→GCB→ORC→Preventive Health Behavior (b) | -0.041 | 0.008 | -0.038 | -0.006 |
| *Indirect effects BHP:* |  |  |  |  |
| BHP→ORC→Preventive Health Behavior | -0.052 | 0.028 | -0.092 | 0.020 |
| BHP→GCB→Preventive Health Behavior | -0.043 | 0.015 | -0.060 | -0.003 |
| BHP→GCB→ORC→Preventive Health Behavior (c) | -0.044 | 0.012 | -0.054 | -0.010 |
| Total effect of AEA on Preventive Health Behavior | -0.008 | 0.117 | -0.266 | 0.184 |
| Total effect of DSP on Preventive Health Behavior | -0.164 | 0.037 | -0.188 | -0.042 |
| Total effect of BHP on Preventive Health Behavior | -0.046 | 0.054 | -0.166 | 0.054 |

*Note:* AEA = Anti-Elitism Attitude; DSP = Demand for Sovereign of the People; BHP = Belief in Homogeneity of People; GCB = Generic Conspiracist Beliefs; ORC = Overconfidence about the Risk of Covid-19

Worth to note, the total effects of the three populist attitudes (table 3), i.e., the algebraic sum of direct and indirect effects (or the effect of populist attitudes on preventive health behaviors when mediators are omitted), give some valuable insights. Indeed, the total effect of AEA on preventive health related behaviors is not significant (*b* = -.008, *se* = .117, *bootstrap 95% C*.*I*. -.266; .184) as well as the total effect of BHP on preventive health behaviors (*b* = -.046, *se* = .054, *bootstrap 95% C*.*I*. -.166; .054). However, the total effect of DSP on preventive health behaviors is still negative and significant (*b* = -.164, *se* = .037, *bootstrap 95% C*.*I*. -.188; -.042). In conclusion, while the total effects of AEA and BHP are suppressed, that of DSP is not.

## Discussion

The goal of our study was to investigate whether populism and beliefs in conspiracy theories, two attitudes sharing a deep distrust toward political and economic elite and experts, may affect the adoption of preventive health behaviors. In particular, in the present study we hypothesized that both populism and conspiracy beliefs may inflate overconfidence about perceived health risk and that this overconfidence may be responsible for the undermining of preventive health behaviors. To verify our hypotheses, we ran an online survey during quarantine and at the beginning of the vaccine release on a national basis and asked participants about their willingness to comply with preventive behaviors against the spread of SARS-CoV-2. Results showed that a populist attitude is associated with a lower compliance with preventive rules. This finding is coherent with previous studies: it has been seen, for example, how the rise of populism was associated with specific attitudes regarding certain preventive behaviors, such as an increase in hesitation to get vaccinated [35]. Moreover, in previous studies, anti-intellectualism (one of the founding elements of the populist attitude) has been associated with the rejection of information provided by experts and scientists [36]. Since preventive behaviors are based precisely on this information (in Italy the Technical-Scientific Committee [CTS] has provided the Government with the necessary information to develop a plan of stringent regulations against the spread of the virus), this could explain why, in our study, populism emerged to be associated with poor compliance with these behaviors. Populist people, in fact, could reject such recommendations due to the lack of trust placed in scientists’ voices.

Likewise, in our study, we found that the tendency to believe in conspiracy theories was also associated with poor adherence to preventive behaviors against Covid-19, a result consistent with what has previously come up in the literature [11, 20, 16]. In fact, the spread of conspiracy theories about viruses such as HIV or about vaccines has already been found to be associated with a lower adherence to preventive behaviors and an increase in hesitation to get vaccinated [37, 15, 38, 39, 14]. The fact that the spread of conspiracy theories represents an obstacle to the adoption of healthy behaviors comes therefore as no surprise if we take into consideration the previous literature [14, 15]. One of the reasons why this happens could be due to a decrease in the perceived severity of the disease, which would lead people to underestimate the risk it can represent for themselves and for others. Indeed, confirming what has been argued so far, our study has shown that the tendency to believe in conspiracy theories is negatively associated with perceived severity, i.e. the belief in conspiracy theories induces an overconfidence about perceived severity of the disease, and this is also coherent with previous works [11, 16].

Overall, therefore, a populist attitude seems to lead to a lower compliance with preventive behaviors (and sometimes to an active and hostile rejection of them) by inflating conspiracy beliefs and the overconfidence about the risk of Covid-19.

## Limitations

Even though the results discussed in our study support our hypotheses and show how political leaders embracing populist themes may affect people overconfidence in the perceived risk for their own health, we would stress the fact that our results are correlational in nature and lack the basis for a causal interpretation of the relationships considered. For this reason, it would be interesting to further explore the results we obtained in a future follow-up and find out whether they can be generalized to other contexts where both populism and conspiracy beliefs may exert their grip. A further theme that needs to be deepened in future studies is related to the relationship between populism and conspiracy beliefs. Literature and results of our study have shown that undoubtedly the two constructs are empirically associated. In our study we placed populism as an antecedent of beliefs in conspiracy beliefs, as we believe that in the Covid-19 epidemic populist leaders have played a crucial role in boosting conspiracist themes to undermine scientific experts. The endorsement of conspiracist themes by populist leaders has inevitably reinforced conspiracy beliefs in populist people. However, the two constructs, populism and conspiracy beliefs, are clearly distinct and it is not excluded that in other contexts the direction of the relationship may be different. Given that in Covid-19 pandemic political leaders have played a pivotal role, future studies should also consider the possible role of partisanship, that is the psychological attachment of an individual towards a specific political party [40]. One of the aspects of partisanship is indeed the tendency to take positions according to what is claimed by the political party with which one identifies the most, without a fair consideration of facts and circumstances. Although Stecula & Pickup’s [20] study has shown that citizens’ positions towards science were not dependent on the construct of partisanship, further investigations on the subject are crucial to extend the results of the aforementioned study (carried out in the American context) also to the Italian setting, in which, without neglecting the Movimento 5 Stelle variable, there would seem to be a clearer correspondence between populist attitudes and the expression of a Right-wing political orientation.

## Notes

### Competing Interest Statement

The authors have declared no competing interest.

### Funding Statement

This study received no funding from any individual or institution.

### Author Declarations

All procedures and measures included in the study were in accordance with the ethical standards approved by the Department of Psychology of Development and Socialization Processes Ethics Committee, which gave approval to our study, and with 1964 Helsinki declaration on ethical standards.

## References

1. Mudde, C., & Kaltwasser, C. R. Exclusionary vs. inclusionary populism: Comparing contemporary Europe and Latin America. Government and opposition, 2013; 48(2): 147–174. https://doi.org/10.1017/gov.2012.11

2. Sutton RM, Douglas KM. 14 Examining the monological nature of conspiracy theories. Power, Politics, and Paranoia: Why People Are Suspicious of Their Leaders. 2014; 29;29: 254–72. https://doi.org/10.1017/CBO9781139565417.018

3. Albertazzi, D., McDonnell, D. Twenty-first century populism, Springer. 2008. http://dx.doi.org/10.1057/9780230592100

4. Hawkins, K. A. Is Chávez populist? Measuring populist discourse in comparative perspective. Comparative Political Studies, 2009; 42(8): 1040–1067. https://doi.org/10.1177%2F0010414009331721

5. Albertazzi, D. Addressing ’the People’: A Comparative Study of the Lega Nord’s and Lega dei Ticinesi’s Political Rhetoric and Styles of Propaganda. Modern Italy, 2007; 12(3): 327–347. https://doi.org/10.1080/13532940701633791

6. Oliver, J. E., & Wood, T. J. Conspiracy theories and the paranoid style (s) of mass opinion. American Journal of Political Science, 2014; 58(4): 952–966. https://www.jstor.org/stable/24363536

7. Hawkins, K. A. Venezuela’s chavismo and populism in comparative perspectives. 2010. Cambridge University Press.

8. Fenster, M. Conspiracy theories: Secrecy and power in American culture. 1999. University of Minnesota Press.

9. Doyle, D. The legitimacy of political institutions: Explaining contemporary populism in Latin America. Comparative Political Studies, 2011; 44(11): 1447–1473. https://doi.org/10.1177%2F0010414011407469

10. Castanho Silva, B., Vegetti, F., & Littvay, L. The elite is up to something: Exploring the relation between populism and belief in conspiracy theories. Swiss political science review, 2017; 23(4): 423–443. https://doi.org/10.1111/spsr.12270

11. Maftei, A., & Holman, A. C. Beliefs in conspiracy theories, intolerance of uncertainty, and moral disengagement during the coronavirus crisis. Ethics & Behavior, 2020; 1–11. https://doi.org/10.1080/10508422.2020.1843171

12. Erceg, N., RuŽojcic, M., & Galić, Z. Misbehaving in the corona crisis: The role of anxiety and unfounded beliefs. Current Psychology, 2020; 1–10. https://doi.org/10.1007/s12144-020-01040-4

13. Fontaine, R. G., Fida, R., Paciello, M., Tisak, M. S., & Caprara, G. V. The mediating role of moral disengagement in the developmental course from peer rejection in adolescence to crime in early adulthood. Psychology, Crime & Law, 2014; 20(1): 1–19. https://psycnet.apa.org/doi/10.1080/1068316X.2012.719622

14. Offit, P. A. Deadly Choices: How the Anti-Vaccine Movement Threatens Us All. 2010. New York: Basic Books.

15. Westergaard, R. P., Beach, M. C., Saha, S., & Jacobs, E. A. Racial/ethnic differences in trust in health care: HIV conspiracy beliefs and vaccine research participation. Journal of general internal medicine, 2014; 29(1): 140–146. https://doi.org/10.1007/s11606-013-2554-6

16. Plohl, N., & Musil, B. Modeling compliance with COVID-19 prevention guidelines: The critical role of trust in science. Psychology, Health & Medicine, 2021; 26(1): 1–12. https://doi.org/10.1080/13548506.2020.1772988

17. Chen, J. Y., Fox, S. A., Cantrell, C. H., Stockdale, S. E., & Kagawa-Singer, M. Health disparities and prevention: racial/ethnic barriers to flu vaccinations. Journal of community health, 2007; 32(1): 5–20. https://doi.org/10.1007/s10900-006-9031-7

18. Lau, J. T., Kim, J. H., Tsui, H. Y., & Griffiths, S. Anticipated and current preventive behaviors in response to an anticipated human-to-human H5N1 epidemic in the Hong Kong Chinese general population. BMC Infectious Diseases, 2007; 7(1): 1–12. https://doi.org/10.1186/1471-2334-7-18

19. Maughan-Brown, B., & Venkataramani, A. S. Accuracy and determinants of perceived HIV risk among young women in South Africa. BMC public health, 2018; 18(1): 1–9. https://doi.org/10.1186/s12889-017-4593-0

20. Stecula, D. A., & Pickup, M. How populism and conservative media fuel conspiracy beliefs about COVID-19 and what it means for COVID-19 behaviors. Research & Politics, 2021; 8(1). https://doi.org/10.1177%2F2053168021993979

21. Schoemann, A. M., Boulton, A. J., & Short, S. D. Determining power and sample size for simple and complex mediation models. Social Psychological and Personality Science, 2017; 8(4): 379–386. https://doi.org/10.1177%2F1948550617715068

22. Jöreskog, K. G., & Sörbom, D. LISREL 8: User’s reference guide. 1996. Chicago, IL: Scientific Software International

23. Yang-Wallentin, F., Jöreskog, K. G., & Luo, H. Confirmatory factor analysis of ordinal variables with misspecified models. Structural Equation Modeling, 2010; 17(3): 392–423. http://dx.doi.org/10.1080/10705511.2010.489003

24. Hu, L., & Bentler, P. M. Cutoff criteria for fit indexes in covariance structure analysis: Conventional criteria versus new alternatives. Structural Equation Modeling, 1999; 6: 1–55. https://doi.org/10.1080/10705519909540118

25. Cronbach, L. J. Coefficient alpha and the internal structure of tests. Psychometrika, 1951; 16(3): 297–334. https://doi.org/10.1007/BF02310555

26. McDonald, R. P. Test theory: A unified treatment. 1999. Mahwah, NJ: Erlbaum.

27. R Core Team R: A language and environment for statistical computing. 2021. R Foundation for Statistical Computing, Vienna, Austria. URL https://www.R-project.org/

28. Revelle, W. psych: Procedures for Personality and Psychological Research, 2021, Northwestern University, Evanston, Illinois, USA, https://CRAN.R-project.org/package=psychVersion=2.1.6

29. Korkmaz S, Goksuluk D, Zararsiz G. MVN: An R Package for Assessing Multivariate Normality. The R Journal. 2014; 6(2): 151–162. http://dx.doi.org/10.32614/RJ-2014-031

30. Rosseel, Y. lavaan: An R Package for Structural Equation Modeling. Journal of Statistical Software, 2012; 48(2): 1-36. URL http://www.jstatsoft.org/v48/i02/

31. Jorgensen, T. D., Pornprasertmanit, S., Schoemann, A. M., & Rosseel, Y. semTools: Useful tools for structural equation modeling. R package version 0.5-5. 2021. Retrieved from https://CRAN.R-project.org/package=semTools

32. MacKinnon, D. P., Lockwood, C. M., & Williams, J. Confidence limits for the indirect effect: Distribution of the product and resampling methods. Multivariate Behavioral Research, 2004; 39: 99–128. https://dx.doi.org/10.1207%2Fs15327906mbr3901_4

33. Schulz, A., Müller, P., Schemer, C., Wirz, D. S., Wettstein, M., & Wirth, W. Measuring populist attitudes on three dimensions. International Journal of Public Opinion Research, 2018; 30(2): 316–326. https://doi.org/10.1093/ijpor/edw037

34. Brotherton, R., French, C. C., and Pickering, A. D. Measuring belief in conspiracy theories: the generic conspiracist beliefs scale. Frontiers in Psychology, 2013; 4: 1–15. http://dx.doi.org/10.3389/fpsyg.2013.00279

35. Kennedy, J. Populist politics and vaccine hesitancy in Western Europe: an analysis of national-level data. European journal of public health, 2019; 29(3): 512–516. https://doi.org/10.1093/eurpub/ckz004

36. Merkley, E. Anti-intellectualism, populism, and motivated resistance to expert consensus. Public Opinion Quarterly, 2020; 84(1): 24–48. https://doi.org/10.1093/poq/nfz053

37. Gaston, G. B., & Alleyne-Green, B. The impact of African Americans’ beliefs about HIV medical care on treatment adherence: a systematic review and recommendations for interventions. AIDS and Behavior, 2013; 17(1): 31–40. https://doi.org/10.1007/s10461-012-0323-x

38. Kata, A. Anti-vaccine activists, Web 2.0, and the postmodern paradigm–An overview of tactics and tropes used online by the anti-vaccination movement. Vaccine, 2012; 30(25): 3778–3789. https://doi.org/10.1016/j.vaccine.2011.11.112

39. Mills, E., Jadad, A. R., Ross, C., & Wilson, K. Systematic review of qualitative studies exploring parental beliefs and attitudes toward childhood vaccination identifies common barriers to vaccination. Journal of clinical epidemiology, 2005; 58(11): 1081–1088. https://doi.org/10.1016/j.jclinepi.2005.09.002

40. Campbell, A., Converse, P. E., Miller, W. E., & Stokes, D. E. The American voter. 1980. University of Chicago Press.

